# The association of diet quality and physical activity with cardiovascular disease and mortality in 85,545 older Australians: A longitudinal study

**DOI:** 10.1101/2023.11.08.23298281

**Authors:** Ding Ding, Joe Van Buskirk, Stephanie Partridge, Philip Clare, Edward Giovannucci, Adrian Bauman, Nicole Freene, Robyn Gallagher, Binh Nguyen

## Abstract

**Background:** A quality diet and an active lifestyle are both important cornerstones of cardiovascular disease (CVD) prevention. However, despite their interlinked effects on metabolic health, the two behaviours are rarely jointly considered, particularly within the context of CVD prevention. We examined the independent, interactive and joint associations of diet and physical activity with CVD hospitalisation, CVD mortality and all-cause mortality.

**Methods:** CVD-free Australian participants aged 45-74 years reported physical activity, diet and sociodemographic and lifestyle characteristics at baseline (2006-09) and follow-up (2012-15), and data were linked to hospitalisation and death registries (31/03/2019 for CVD hospitalisation and all-cause mortality and 08/12/2017 for CVD mortality). Diet quality was categorised as low, medium and high based on meeting dietary recommendations. Physical activity was operationalised as 1) total moderate-to-vigorous physical activity (MVPA) as per guidelines and 2) the composition of MVPA as the ratio of vigorous-intensity physical activity (VPA) to total MVPA. We used a left-truncated cause-specific Cox proportional hazards model using time-varying covariates.

**Results:** During a median of 10.7 years of follow-up, 6,581 participants were admitted to the hospital for CVD and 6,586 died from all causes (879 from CVD during 9.3 years). A high-quality diet was associated with a 17% lower risk of all-cause mortality than low-quality, and the highest MVPA category (compared with the lowest) was associated with 44% and 48% lower risks of CVD and all-cause mortality. Multiplicative interactions between diet and physical activity were non-significant. For all outcomes, the lowest risk combinations involved a high-quality diet and the highest MVPA categories. Accounting for total MVPA, some VPA was associated with further risk reduction of CVD hospitalisation and all-cause mortality.

**Conclusion:** For CVD prevention and longevity, one should adhere to both a healthy diet and an active lifestyle and incorporate some VPA when possible.

## Introduction

Cardiovascular disease (CVD) is the leading cause of global disease burden, responsible for premature mortality, substantial healthcare costs, reduced productivity and quality of life.^1^ In Australia, CVD was responsible for more than 5% of all hospitalisations and 25% of all deaths in 2020-21.^2^ Many CVDs, such as ischemic heart disease^3^ and stroke^4^ are largely preventable through a healthy lifestyle. Both a quality diet and sufficient physical activity are recommended by health authorities^5^ ^6^ as important cornerstones of an ‘optimal lifestyle’ for CVD prevention.

For decades, researchers have argued that diet and physical activity be jointly examined in epidemiological studies, considering that the two are inherently linked through energy balance and metabolic homeostasis.^7^ ^8^ However, to date, only a small number of studies have examined interactive or joint associations between diet and physical activity,^9–12^ and even fewer have explored CVD outcomes.^11^ ^13^ ^14^ Furthermore, considering that diet and physical activity are likely to vary over time, only one study to our knowledge examined measures at more than one point in time.^12^

In this study, we examined the independent, interactive, and joint associations of diet and physical activity with CVD hospitalisation and CVD mortality (primary outcomes) and all-cause mortality (secondary outcome) in a large cohort of middle-aged and older Australian adults, the 45 and Up study.

## Methods

### Sampling and procedures

The 45 and Up study is a large population-based prospective cohort study of middle-aged and older adults from New South Wales (NSW), the most populous state in Australia. Details about the study are documented elsewhere.^15^ Briefly, in 2006-2009, NSW adults aged 45 and older were randomly sampled from the database of Medicare, Australia’s publicly funded universal health care insurance scheme. The database includes all citizens and permanent residents and some temporary residents and refugees. To participate, contacted individuals completed and returned the questionnaire, together with a signed consent form (including consent for long-term follow-up by linkage to administrative data such as hospital records), in a pre-paid envelope. The baseline sample (n=267,153) accounted for around 10% of the whole population of NSW aged 45 and over at the time, and around 18% of those invited to participate.^15^ In 2012-2015, 246,306 participants were invited to complete a follow-up survey. Of those, 142,548 (58%) responded (Supplementary Figure 1).

### Measurement

Questionnaires for the baseline and follow-up surveys are available at http://www.45andUp.org.au. All independent variables and covariates were measured at both time points using the same instruments.

#### Exposures

Physical activity was measured using the Active Australia survey^16^ which has reliability and validity deemed to be acceptable^17^ ^18^ and comparable with commonly used physical activity surveys.^19^ Participants were asked to recall the total time of walking (for recreation, exercise or getting to and from places that lasted at least for 10 minutes), moderate-intensity (with examples provided such as “gentle swimming, social tennis”) and vigorous-intensity activity (VPA; defined as activities “that made you breathe harder or puff and pant, like jogging, cycling, aerobics, competitive tennis, but not household chores or gardening”) in the past week. Two physical activity measures were created. First, total moderate-to-vigorous intensity physical activity (MVPA) was calculated as the sum of walking, moderate-intensity physical activity, and VPA (weighted by 2 to account for its higher energy) as per survey protocol.^16^ Total MVPA was further categorised into 0-9, 10-149, 150-299, 300-800, 900+ minutes/week based on distribution of status regarding meeting physical activity guidelines. Second, considering that previous studies have indicated potential added benefits conveyed by VPA, while accounting for the overall activity volume, we created an additional variable denoting the proportion of VPA to total MVPA (VPA% thereafter), following the practice of previous studies.^20^ ^21^ We further categorised VPA% as no VPA, >0-20% VPA, >20-50% VPA, and 50% VPA, based on spline models (Supplementary Figure 2) and a previous study from the same cohort.^20^

Diet quality was assessed using a repeated short questionnaire which included dietary frequency measures of several food items, such as red meat, poultry, and cheese, as well as daily serves of fruit and vegetables. To accommodate CVD outcomes, we modified a previously developed diet quality score^22^ to include fruit, vegetables, fish, red meat and processed meat. The selection and scoring of diet items are based on the Australian Dietary Guidelines^23^ and a Heart Foundation Position Statement.^24^ Each diet item score ranged from 0 (the lowest quality) to 2 (the highest), which contributed to the total diet quality score ranging from 0 to 10 (Supplementary Table 1). We further categorised the total score into low (0-4), medium (5-7) and high (8-10) based on data distribution and the functional form of the association between diet quality score and CVD outcomes (Supplementary Figure 2).

Specifically, univariate models were fitted for the association between diet score and all outcomes, visually inspecting each category’s association to test for ordered grouping in the data to determine cut points.

### Outcomes

The primary outcomes were defined as 1) the first CVD hospitalisation (ascertained from NSW Admitted Patient Data Collection) following the completion of the baseline questionnaire with a primary diagnosis of ischemic heart disease (ICD-10 codes I20-25), heart failure (I50) or stroke (I60-I64);^25^ and 2) CVD mortality defined as death by CVD (I00-I99) as the primary cause ascertained from the NSW Cause of Death Unit Record File. Our decision of focusing on ischemic heart disease, stroke and heart failure for CVD hospitalisation and broader definitions of CVD for CVD mortality is in line with the approach taken by the 2018 US Physical Activity Guidelines Advisory Committee.^26^ The secondary outcome was all-cause mortality ascertained from the NSW Registry of Births Deaths & Marriages. In all analyses, participants contributed person-years from the baseline until event or censoring (31/03/2019 for CVD hospitalisation and all-cause mortality, and 08/12/2017 for CVD mortality), whichever occurred first (for Censoring proportion, see Supplementary Table 2).

### Covariates

Covariates were selected based on an a-priori-developed directed acyclic graph, a conceptual representation for mapping assumptions regarding variables involved in a causal research question (Supplementary Figure 3).^27^ The following potential confounders were selected for adjustment: age, sex, marital status (single, married/de facto, widowed, divorce/separated), country of birth (Australia vs overseas), highest educational attainment (no school certificate, school certificate, higher school certificate, trade/apprenticeship/diploma, university degree or higher), private health insurance (yes vs no, as an additional socioeconomic status indicator), area-level socioeconomic status, measured by the Index of Relative Socio-economic Disadvantage (IRSD),^28^ operationalized as quintiles, remoteness, measured by Accessibility/Remoteness Index of Australia (ARIA+),^29^ smoking (never, former, current), alcohol (0, 1-14, 14+ serves per week), family history of CVD (yes vs no, defined by known CVD in first-degree family members), body mass index (BMI; categorised as normal weight [18.5-24.9 kg/m^2^], overweight [25-29.9 kg/m^2^], obese [30+ kg/m^2^]), self-reported diagnosis of diabetes and reported diagnosis of hypertension (yes vs no).

BMI, diagnosis of diabetes and hypertension could be either a precursor or outcome of exposures. ^11^ For the main analysis, we consider BMI, diabetes, and hypertension to be primarily confounders and we adjust for them in our main analysis.

### Statistical analysis

We have pre-registered our analytical protocol with Open Science Framework (OSF) https://doi.org/10.17605/OSF.IO/5VQEH. However, we could not perform all tests registered due to hardware constraints. The reporting of our study adhered to the Strengthening the Reporting of Observational Studies in Epidemiology (STROBE) guidelines.

The analytical sample consisted of participants aged 45 to 74 years at baseline (as our main outcome of interest was CVD at an earlier age),^20^ without a CVD diagnosis prior to baseline (based on reported medical diagnosis, heart surgery and recent medication for CVD in baseline questionnaire and retrospective linkage data to NSW Admitted Patient Data Collection for up to five years prior to baseline), who completed both waves of the questionnaire, as well as those who completed the baseline questionnaire only and experienced the outcome before the end of recruitment for wave 2. We further excluded participants who needed regular help with daily activities and those who were ‘limited a lot’ to walk 100 meters as well as those who were underweight (BMI<18.5 kg/m^2^).

We examined the relationship between exposures and outcomes using a left-truncated cause-specific Cox proportional hazards models using time-varying covariates and age as the underlying time scale.^30^ We assessed any violation of the proportional hazards assumption by using Schoenfield residuals and stratified covariates if necessary until the assumption was met. We calculated hazard ratios (HR) and their 95% confidence intervals (CI) to present the instantaneous, proportional hazard of each outcome. To avoid informative censoring due to drop out, we weighted participants according to their inverse-probability of appearing in wave 2, with probabilities calculated using logistic regression and adjusted for all covariates included in the final model.^31^

To examine the independent and joint associations between physical activity and diet, we built several models. For independent associations, both diet and physical activity (MVPA and VPA% in separate models) were included in the same model while adjusted for all covariates listed above, and mutually adjusted for each other. The VPA% model further adjusted for total MVPA and was conducted among the subsample of the participants with at least 10 minutes of MVPA (to avoid zero or nearly zero denominator). We included a diet × physical activity interaction term in the fully adjusted model and performed a Wald’s test. For joint associations, we created 15 (3×5) mutually exclusive diet and MVPA linear combinations, and 12 (3×4) mutually exclusive diet and VPA% linear combinations and used them as the exposures. This allows for comparing different exposure categories against the same reference. We used the combination for no MVPA and low diet quality score (theoretically highest risk combination) as the reference for the MVPA analysis and the combination of 0% VPA and low diet quality score for VPA% analysis. Data were analysed in R 4.2.2 (R Core Team, 2017) and a 2-tailed alpha was set at 0.05.

We conducted two sensitivity analyses. First, to account for reverse causation by pre-existing disease, we reran all analyses excluding the first two years of follow-up as ‘washout’ period. We further repeated this sensitivity analysis using a five-year ‘washout’ period for CVD and all-cause mortality because mortality outcomes may take longer to manifest. Second, as mentioned, BMI, diabetes and hypertension could be confounders or mediators. For the sensitivity analysis, we re-considered them as mediators and therefore did not adjust for them in the model.

#### Missing data

The survey included missing data due to both incomplete surveys and loss to follow-up. Further detail on the amount of missing data in each variable is included in Supplementary Table 3. Missing data were handled using multiple imputation using a random forest algorithm, and with the number of imputations determined based on Monte Carlo error linked to the amount of missing information.^32^ Specifically, 20 multiple imputations were run on each outcome-specific dataset, each with a maximum of 50 iterations.

## Results

The final study sample included 85,545 participants aged 45 to 74 years, without a CVD diagnosis or severe physical limitations and that were not underweight at baseline (Supplementary Figure 1). During a median of 10.7 (interquartile range [IQR]=10.5 to 11.1) years of follow-up, 6,576 participants were admitted to the hospital for CVD, 6,581 died from all causes. Specifically, during a median of 9.3 (IQR=9.2 to 9.8) years of follow up, 876 died from CVD causes.

### Baseline characteristics

Participants characteristics at baseline by MVPA and diet quality are presented in Table 1 (and by VPA%, see Supplementary Table 4). Overall, participants had a mean age of 58.2 (SD=7.55) (median=57.6, IQR=12). More than half of the participants were women (55.4%), 32.2% had a university degree or higher, 81.0% were married/de facto, 22.1% were born overseas, and 51.9% resided in major cities. Some 32.8% participants were smoking at baseline, 16.1% reported drinking more than 14 serves/week of alcohol, and 40.7% were overweight and 20.5% obese. More than half of the participants reported having a family history of CVD (56.1%), 29.0% reported a diagnosis of hypertension at baseline and 5.3% reported diabetes. Overall, they reported a mean of 630 (SD=591) minutes of weekly participation in MVPA (median=460, IQR=615); 16% reported meeting minimal recommendations of 150-299 minutes of MVPA per week and 67.7% reported 300 or more minutes per week. Among those who reported doing any MVPA, 51.8% reported some VPA. The median diet quality score is 5 (IQR=2) with 9.0%, 59.8%, and 31.2% classified as low, medium, and high diet quality, respectively.

**Table 1.** Descriptive statistics of the study sample at baseline (n=85,545, 2005-2019)

|  | Physical activity (min/week) |  |  |  |  | Diet quality score |  |  |
| --- | --- | --- | --- | --- | --- | --- | --- | --- |
|  | <10<br>(N=2806) | 10-149<br>(N=10979) | 150-299<br>(N=13750) | 300-899<br>(N=23541) | 900+<br>(N=34469) | Low<br>(N=7697) | Medium<br>(N=51128) | High<br>(N=26720) |
| **Age:** Mean (SD) years | 58.0 (7.60) | 57.7 (7.59) | 57.4 (7.47) | 57.6 (7.44) | 59.1 (7.55) | 57.7 (7.68) | 58.1 (7.61) | 58.5 (7.36) |
| **Female** | 1434 (51.1%) | 5666 (51.6%) | 7540 (54.8%) | 13338 (56.7%) | 19081 (55.4%) | 2533 (32.9%) | 25895 (50.6%) | 18631 (69.7%) |
| **Education** |  |  |  |  |  |  |  |  |
| No school certificate | 407 (14.5%) | 835 (7.6%) | 737 (5.4%) | 1216 (5.2%) | 2279 (6.6%) | 666 (8.7%) | 3298 (6.5%) | 1510 (5.7%) |
| School certificate only | 688 (24.5%) | 2186 (19.9%) | 2462 (17.9%) | 4104 (17.4%) | 6776 (19.7%) | 1598 (20.8%) | 9715 (19.0%) | 4903 (18.3%) |
| Higher school certificate only | 292 (10.4%) | 1117 (10.2%) | 1311 (9.5%) | 2160 (9.2%) | 3324 (9.6%) | 864 (11.2%) | 4971 (9.7%) | 2369 (8.9%) |
| Trade/apprenticeship/diploma | 865 (30.8%) | 3533 (32.2%) | 4276 (31.1%) | 7438 (31.6%) | 12034 (34.9%) | 2803 (36.4%) | 16944 (33.1%) | 8399 (31.4%) |
| University or higher | 554 (19.7%) | 3308 (30.1%) | 4964 (36.1%) | 8623 (36.6%) | 10056 (29.2%) | 1766 (22.9%) | 16200 (31.7%) | 9539 (35.7%) |
| **Area-level socioeconomic disadvantage** |  |  |  |  |  |  |  |  |
| Quintile 1 (most disadvantaged) | 661 (23.6%) | 2007 (18.3%) | 2152 (15.7%) | 3621 (15.4%) | 5724 (16.6%) | 1488 (19.3%) | 8401 (16.4%) | 4276 (16.0%) |
| Quintile 2 | 615 (21.9%) | 2241 (20.4%) | 2679 (19.5%) | 4430 (18.8%) | 7128 (20.7%) | 1732 (22.5%) | 10164 (19.9%) | 5197 (19.4%) |
| Quintile 3 | 578 (20.6%) | 2108 (19.2%) | 2642 (19.2%) | 4407 (18.7%) | 6811 (19.8%) | 1492 (19.4%) | 9895 (19.4%) | 5159 (19.3%) |
| Quintile 4 | 484 (17.2%) | 2021 (18.4%) | 2666 (19.4%) | 4516 (19.2%) | 6458 (18.7%) | 1407 (18.3%) | 9755 (19.1%) | 4983 (18.6%) |
| Quintile 5 (least disadvantaged) | 468 (16.7%) | 2602 (23.7%) | 3611 (26.3%) | 6567 (27.9%) | 8348 (24.2%) | 1578 (20.5%) | 12913 (25.3%) | 7105 (26.6%) |
| **Marital status** |  |  |  |  |  |  |  |  |
| Single | 187 (6.7%) | 740 (6.7%) | 887 (6.5%) | 1363 (5.8%) | 1901 (5.5%) | 518 (6.7%) | 2815 (5.5%) | 1745 (6.5%) |
| Married/de facto | 2215 (78.9%) | 8745 (79.7%) | 11070 (80.5%) | 19168 (81.4%) | 28105 (81.5%) | 6293 (81.8%) | 42211 (82.6%) | 20799 (77.8%) |
| Widowed | 93 (3.3%) | 390 (3.6%) | 468 (3.4%) | 813 (3.5%) | 1271 (3.7%) | 203 (2.6%) | 1612 (3.2%) | 1220 (4.6%) |
| Divorced/separated | 311 (11.1%) | 1104 (10.1%) | 1325 (9.6%) | 2197 (9.3%) | 3192 (9.3%) | 683 (8.9%) | 4490 (8.8%) | 2956 (11.1%) |
| **Born overseas** | 592 (21.1%) | 2414 (22.0%) | 3002 (21.8%) | 5156 (21.9%) | 7777 (22.6%) | 1242 (16.1%) | 11048 (21.6%) | 6651 (24.9%) |
| **Residential location** |  |  |  |  |  |  |  |  |
| Major Cities | 1368 (48.8%) | 5988 (54.5%) | 7658 (55.7%) | 12900 (54.8%) | 16424 (47.6%) | 3613 (46.9%) | 26349 (51.5%) | 14376 (53.8%) |
| Inner Regional | 1050 (37.4%) | 3773 (34.4%) | 4705 (34.2%) | 8335 (35.4%) | 13620 (39.5%) | 2891 (37.6%) | 18842 (36.9%) | 9750 (36.5%) |
| Outer Regional/Remote | 388 (13.8%) | 1218 (11.1%) | 1387 (10.1%) | 2306 (9.8%) | 4425 (12.8%) | 1193 (15.5%) | 5937 (11.6%) | 2594 (9.7%) |
| **Smoking status** |  |  |  |  |  |  |  |  |
| Never | 1518 (54.1%) | 6667 (60.7%) | 8554 (62.2%) | 14385 (61.1%) | 20132 (58.4%) | 3902 (50.7%) | 30400 (59.5%) | 16954 (63.5%) |
| Previous | 904 (32.2%) | 3384 (30.8%) | 4266 (31.0%) | 7723 (32.8%) | 11819 (34.3%) | 2715 (35.3%) | 16905 (33.1%) | 8476 (31.7%) |
| Current | 384 (13.7%) | 928 (8.5%) | 930 (6.8%) | 1433 (6.1%) | 2518 (7.3%) | 1080 (14.0%) | 3823 (7.5%) | 1290 (4.8%) |
| **Alcohol consumption** |  |  |  |  |  |  |  |  |
| 0 serves/week | 1068 (38.1%) | 3455 (31.5%) | 3762 (27.4%) | 5695 (24.2%) | 8709 (25.3%) | 1808 (23.5%) | 12920 (25.3%) | 7961 (29.8%) |
| 1-14 serves/week | 1274 (45.4%) | 5921 (53.9%) | 7927 (57.7%) | 14219 (60.4%) | 19700 (57.2%) | 3719 (48.3%) | 29278 (57.3%) | 16044 (60.0%) |
| >14 serves/week | 464 (16.5%) | 1603 (14.6%) | 2061 (15.0%) | 3627 (15.4%) | 6060 (17.6%) | 2170 (28.2%) | 8930 (17.5%) | 2715 (10.2%) |
| **Body mass index (kg/m<sup>2</sup>)** |  |  |  |  |  |  |  |  |
| Normal weight (18.5-24.9) | 738 (26.3%) | 3381 (30.8%) | 4949 (36.0%) | 9384 (39.9%) | 14789 (42.9%) | 2471 (32.1%) | 18758 (36.7%) | 12012 (45.0%) |
| Overweight (25-29.9) | 1050 (37.4%) | 4431 (40.4%) | 5637 (41.0%) | 9722 (41.3%) | 13964 (40.5%) | 3368 (43.8%) | 21339 (41.7%) | 10097 (37.8%) |
| Obese (30+) | 1018 (36.3%) | 3167 (28.8%) | 3164 (23.0%) | 4435 (18.8%) | 5716 (16.6%) | 1858 (24.1%) | 11031 (21.6%) | 4611 (17.3%) |
| **Family history of CVD** | 1502 (53.5%) | 5897 (53.7%) | 7700 (56.0%) | 13307 (56.5%) | 19620 (56.9%) | 4112 (53.4%) | 28287 (55.3%) | 15627 (58.5%) |
| **Diagnosis of hypertension** | 937 (33.4%) | 3459 (31.5%) | 4103 (29.8%) | 6604 (28.1%) | 9686 (28.1%) | 2322 (30.2%) | 15102 (29.5%) | 7365 (27.6%) |
| **Diagnosis of diabetes** | 227 (8.1%) | 827 (7.5%) | 750 (5.5%) | 1111 (4.7%) | 1648 (4.8%) | 406 (5.3%) | 2829 (5.5%) | 1328 (5.0%) |

Data are presented as n (%) unless otherwise specified.
Abbreviations: CVD=cardiovascular disease, SD=standard deviation.
<sup>1</sup>Physical activity minutes/week calculated as walking + moderate-intensity + vigorous-intensity physical activity ×2 using the Active Australia Questionnaire.
<sup>2</sup>The Diet quality score was developed based on reported consumption of fruit, vegetable, fish, processed meat and red meat. An overall score ranged from 0 (lowest quality) to 10 (highest quality) and was recoded into 0-4 (low), 5-7 (medium) and 8-10 (high).

In most cases, those who were more active (both in terms of MVPA and VPA%) were more likely to have a university degree, and less likely to be smokers or obese. Those with high dietary quality score were more likely to be women and less likely to be current smokers, obese or consume >14 serves/week of alcohol.

### Independent associations of diet and physical activity with outcomes

MVPA, VPA% and diet quality score were associated with all outcomes in unadjusted analyses and the associations were substantially attenuated in adjusted analyses (Table 2). In adjusted models, diet score was only associated with all-cause mortality, where high-quality diet was associated with a 17% reduced risk compared with low-quality. MVPA was significantly associated with both CVD mortality and all-cause mortality in a dose-response fashion where the highest level of MVPA was associated with around half of the risk of CVD and all-cause mortality compared with those who reported the lowest level of MVPA. With respect to VPA, while accounting for total MVPA, having 20-50% and >50% of the MVPA from VPA was significantly associated with an 8% and 11% risk reduction of CVD hospitalisation compared with no VPA, while any non-zero VPA was significantly associated with around 20% of risk reduction for all-cause mortality compared with no VPA.

**Table 2.**
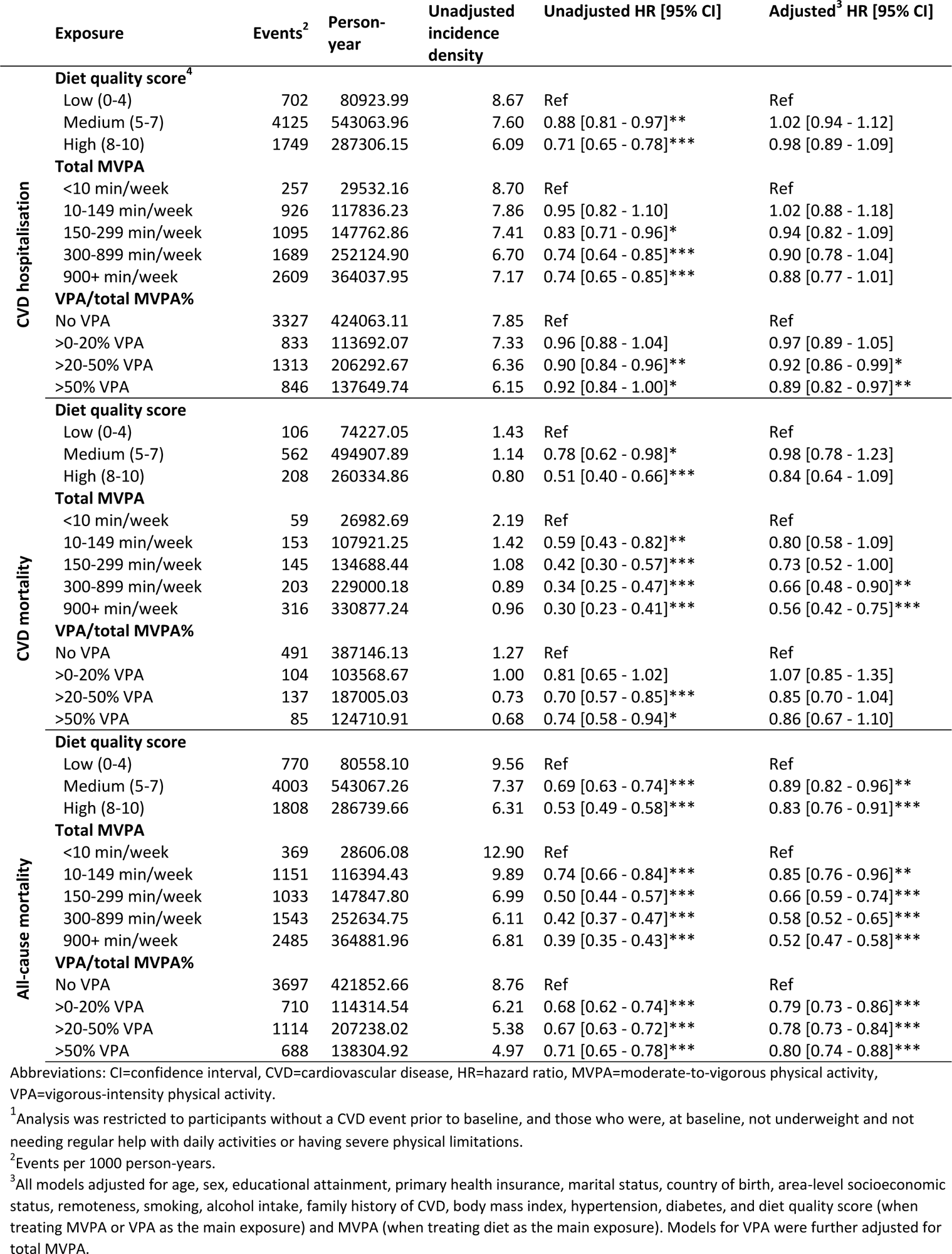
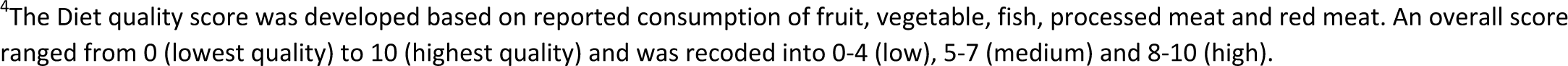
Associations of moderate-to-vigorous physical activity (MVPA), vigorous-intensity physical activity (VPA) and diet quality score with cardiovascular disease (CVD) hospitalisation, CVD mortality and all-cause mortality (n=85,545)^1^.

### Joint associations of diet and physical activity with outcomes

We did not find statistically significant multiplicative interactions between diet and physical activity, suggesting that the association between physical activity and outcomes were similar across diet quality strata, and vice versa. For CVD hospitalisation, nearly none of the combinations tested was statistically different from the reference category (Figures 1 and 2). For CVD mortality, all combinations involving the top two categories of MVPA (corresponding to 150+ minutes/week of MVPA) were associated with significantly lower risk. For all-cause mortality, any combination had significantly lower risk compared with the reference category. Among those that reported at least 10 minutes/week of MVPA, combinations involving any none-zero VPA values were associated with lower risk of all-cause mortality compared with the reference category of zero VPA and low-quality diet.

**Figure 1.**
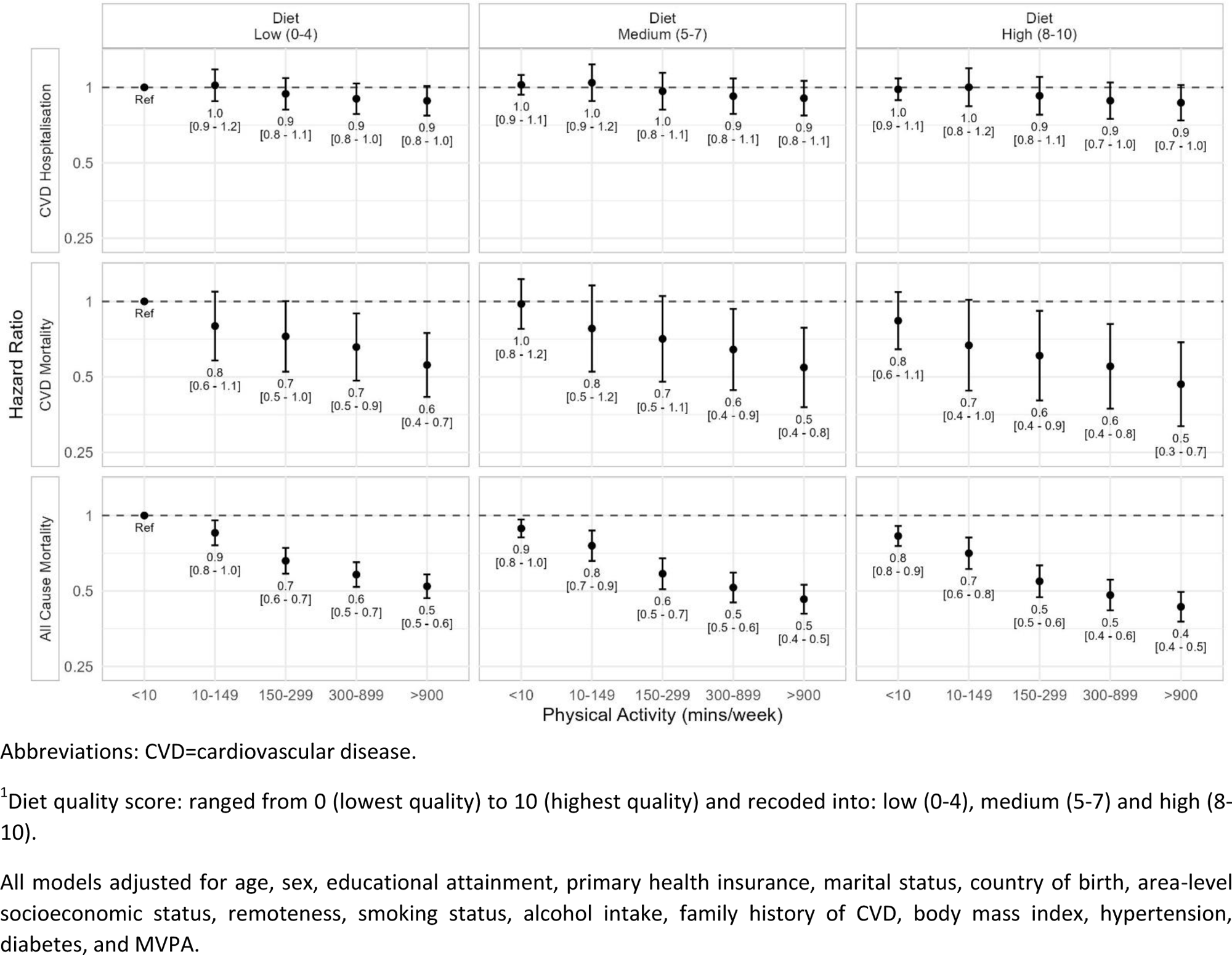
Combinations of moderate-to-vigorous physical activity (MVPA) quartiles and diet quality score^1^ and CVD hospitalisation, CVD mortality and all-cause mortality (n=85,545)

**Figure 2.**
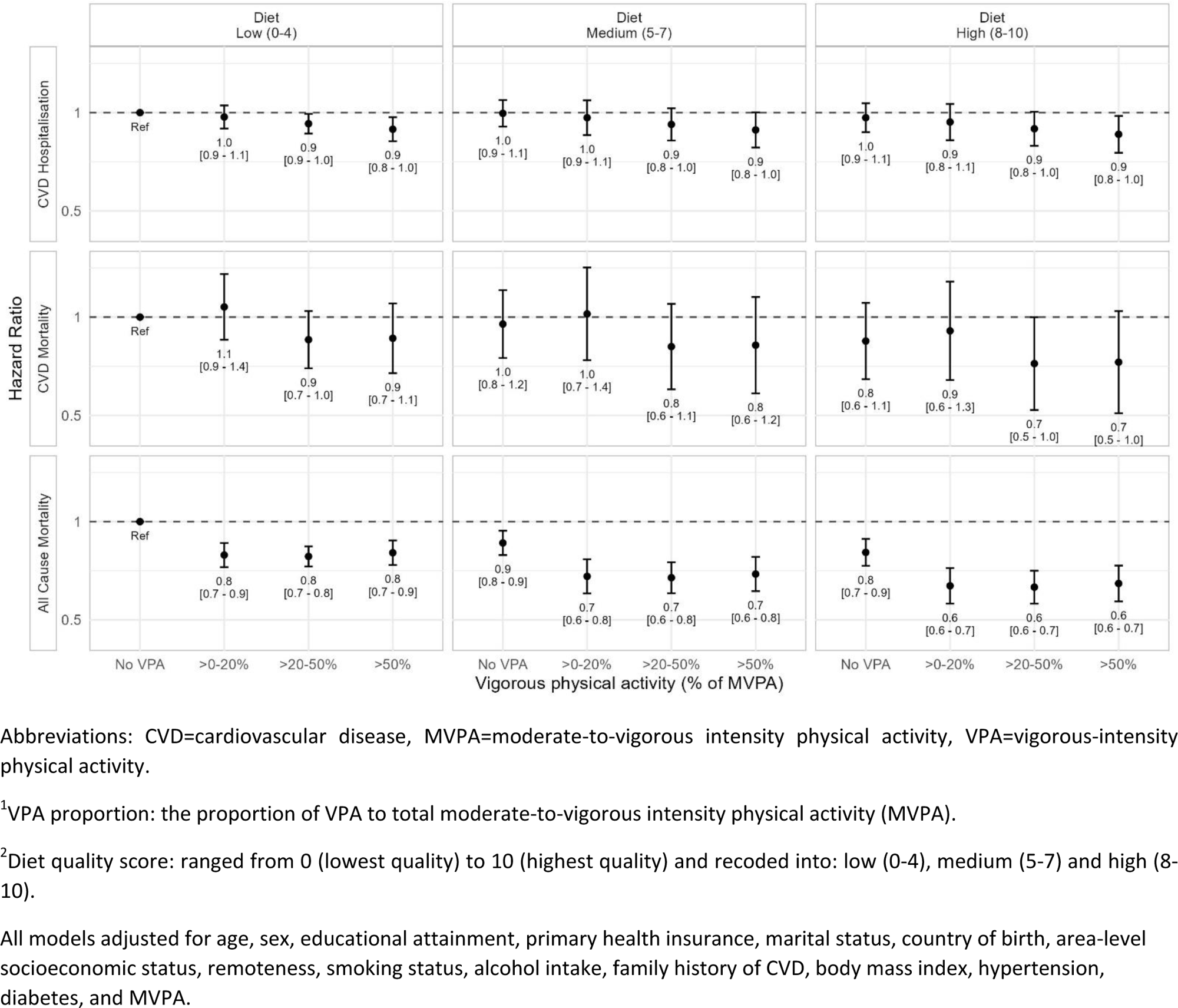
Combinations of vigorous-intensity physical activity (VPA) proportion^1^ and diet quality score^2^ and CVD hospitalisation, CVD mortality and all-cause mortality among those who engaged in at least 10 minutes of physical activity a week (n=82,733)

All sensitivity analyses revealed very similar patterns of associations with minimal differences in the magnitude or significance of associations (Supplementary Tables 5-7; Supplementary Figures 4-12).

## Discussion

Diet and physical activity are both critical modifiable risk factors for CVD. Health authorities, such as the American Heart Association^33^ and Heart Foundation Australia,^24^ recommend an active lifestyle and a healthy diet, often characterised by plenty of fruit, vegetables, wholegrains, fish/seafood, and healthy fats. Based on repeated surveys and linked administrative health records, this study is one of the first to examine the joint association of diet and physical activity on CVD outcomes. Our findings reinforce an important public health message that adhering to both a quality diet and sufficient physical activity is key to CVD prevention and long-term survival.

Our study found inverse associations between physical activity and CVD and all-cause mortality. Specifically, MVPA, particularly at the recommended levels of meeting guidelines,^34^, was associated with significantly lower risk (27-44% for CVD mortality and 34-48% for all-cause mortality). While accounting for total MVPA, VPA% seemed to convey additional benefits, particularly in terms of CVD hospitalisation and all-cause mortality. This finding corroborates existing evidence on the relationship between physical activity and mortality.^21^ ^34^ ^35^ Surprisingly, we did not find a significant association between physical activity and CVD hospital admissions, despite a well-established strong association between physical activity and CVD.^36^ This may be because CVD hospital admission is an imperfect proxy for CVD incidence as it could not capture CVD diagnosis outside of the hospital. This limitation has been acknowledged by previous work using the 45 and Up data.^37^

We found that those with the highest diet quality score had a 17% lower risk of all-cause mortality compared with those with the lowest score. However, we did not find a significant association between diet quality and both CVD outcomes. The lack of association contradicts the overall evidence on dietary quality score and CVD outcomes.^38^ The current diet quality score was based on fruit, vegetables, fish, red meat and processed meat, all of which are important food components for CVD prevention.^23^ ^24^ However, the short dietary questions, such as the one used in the 45 and Up study, may not be comprehensive enough to capture other important dietary components for cardiovascular health, such as nuts. Participants may also have difficulty recalling their dietary patterns without a clearly defined recall period. Questions were not asked consistently across food items (fruit and vegetables were asked in terms of serves per day while the rest of the items included were asked in terms of weekly frequency). Such measurement errors are likely to bias the association towards the null.^40^ A recent meta-analysis on diet quality score based on comprehensive dietary questionnaires found that those with the highest quality diet had around 20% lower risk of all-cause mortality, CVD incidence or mortality compared with those with the lowest quality diet.^41^ The associations observed in our study were of similar magnitude for CVD and all-cause mortality (albeit not significant for CVD mortality) but of much smaller magnitude for CVD hospitalisation.

For the analysis of the joint physical activity and dietary profiles, we did not find any significant multiplicative interactions. Our findings suggest that the lowest risk of CVD and all-cause mortality corresponded to the highest diet quality combined with the highest levels of physical activity. For example, the combination of high-quality diet score and >300 minutes/week of MVPA was associated with around 40-50% lower risk of CVD mortality, and around 50-60% lower risk of all-cause mortality. These findings reinforce the importance of following a healthy diet and being physically active for chronic disease prevention and longevity and align with findings from previous evidence. Several studies have reported greater risk reductions for all-cause mortality with higher physical activity levels combined with better adherence to the Mediterranean diet such as in the Seguimiento Universidad de Navarra (SUN) cohort,^9^ or better diet quality in the Golestan cohort.^14^ However, in the Melbourne Collaborative Cohort Study, increasing adherence to the Mediterranean diet did not appear to provide much added benefit.^12^ Among the few studies examining the joint associations of physical and diet with cardiovascular outcomes, larger reductions in risks of incident CVD and CVD mortality^13^ were reported when higher levels of physical activity were combined with better adherence to the Mediterranean diet^13^ or better diet quality.^14^

### Strengths and limitations

This study is among the first to examine the joint association of physical activity and diet with CVD outcomes. We conducted a comprehensive analysis based on reported physical activity and diet with more than 10 years of follow-up. We applied time-varying covariates approach to consider exposures at more than one time point and applied censoring weights to account for information censoring due to drop out. We have also conducted several sensitivity analyses which confirmed the robustness of our findings. However, several limitations applied. First, both physical activity and diet quality were self-reported and subject to measurement biases. Future studies should consider using accelerometers, dietary recall or comprehensive food frequency questionnaires^42^ for more valid measures of physical activity and diet. Second, CVD hospital admission is not an ideal measure for CVD incidence as it may have missed incident CVD outside of the hospital setting. Third, despite our best efforts to select potential confounders based on directed acyclic graphs, our analysis may still be subject to residual confounding from unmeasured or insufficiently measured confounders. Fourth, our hardware constraints limited our plans to test for the additive interactions between diet and physical activity. Finally, the 45 and Up study had a participation rate of around 18% and tends to overrepresent those who are healthier. For example, in this study, participants reported much higher levels of physical activity compared with general adults in NSW using a comparable measure.^43^ However, despite different prevalence estimates, a study comparing the 45 and Up cohort with a population representative sample in NSW found similar magnitude of associations between risk factors and health-related outcomes.^44^ Furthermore, despite the high levels of physical activity in the 45 and Up participants, associations between physical activity and various outcomes observed in this cohort are still comparable with those from other studies.^45^

### Conclusions

Adhering to both high-quality diet and higher levels of physical activity is associated with the lowest risk of CVD mortality and all-cause mortality, compared with other combinations of the two behaviours. Doing some VPA, such as around 20-50% of total MVPA, seems to offer additional benefits to the combinations of behaviours. Diet and physical activity should be jointly considered in research and promoted as important strategies for CVD prevention.

## Declarations

### Ethical approval and consent to participate

The study was approved by the New South Wales Population and Health Services Research Ethics Committee (reference No. 2010/05/234). Participants provided written consent form (including consent for long-term follow-up by linkage to administrative data such as hospital records) before participating in the study.

### Consent for publication

N/A

### Availability of data and materials

Data can be accessible upon application to the data custodian (the Sax Institute) with a fee.

### Authors’ contributions

DD conceptualised the study, DD and BN conducted literature search, JVB managed and analysed the data, PC advised on data analysis, DD drafted the paper with the support of JVB and BN. All authors contributed to interpreting data and critically reviewing the paper, and all approved the final version of the paper.

## Data Availability

Data are available from data custodians upon request and with a fee

## Acknowledgement

This research was completed using data collected through the 45 and Up Study (www.saxinstitute.org.au). The 45 and Up Study is managed by the Sax Institute in collaboration with major partner Cancer Council NSW and partners the Heart Foundation and the NSW Ministry of Health. We thank the many thousands of people participating in the 45 and Up Study.

## References

1. Roth GA, Mensah GA, Johnson CO, et al. Global Burden of Cardiovascular Diseases and Risk Factors, 1990–2019. J Am Coll Cardiol 2020;76(25):2982–3021. doi: doi:10.1016/j.jacc.2020.11.010

2. Australian institute of Health and Welfare (AIWH). Heart, stroke and vascular disease: Australian facts 2023 [Available from: https://www.aihw.gov.au/reports/heart-stroke-vascular-diseases/hsvd-facts/contents/all-heart-stroke-and-vascular-disease#deaths.

3. Gupta R, Wood DA. Primary prevention of ischaemic heart disease: populations, individuals, and health professionals. Lancet 2019;394(10199):685–96. doi: 10.1016/S0140-6736(19)31893-8

4. Hankey GJ. Population Impact of Potentially Modifiable Risk Factors for Stroke. Stroke 2020;51(3):719–28. doi: doi:10.1161/STROKEAHA.119.024154

5. Arnett DK, Blumenthal RS, Albert MA, et al. 2019 ACC/AHA Guideline on the Primary Prevention of Cardiovascular Disease: A Report of the American College of Cardiology/American Heart Association Task Force on Clinical Practice Guidelines. Circulation 2019;140(11):e596–e646. doi: doi:10.1161/CIR.0000000000000678

6. Visseren FLJ, Mach F, Smulders YM, et al. 2021 ESC Guidelines on cardiovascular disease prevention in clinical practice: Developed by the Task Force for cardiovascular disease prevention in clinical practice with representatives of the European Society of Cardiology and 12 medical societies With the special contribution of the European Association of Preventive Cardiology (EAPC). Eur Heart J 2021;42(34):3227–337. doi: 10.1093/eurheartj/ehab484

7. Blair SN, Horton E, Leon AS, et al. Physical activity, nutrition, and chronic disease. Med Sci Sports Exerc 1996;28(3)

8. Baranowski T. Why combine diet and physical activity in the same international research society? Int J Behav Nutr Phys Act 2004;1(1):2. doi: 10.1186/1479-5868-1-2 [published Online First: 2004/06/03]

9. Alvarez-Alvarez I, Zazpe I, Pérez de Rojas J, et al. Mediterranean diet, physical activity and their combined effect on all-cause mortality: The Seguimiento Universidad de Navarra (SUN) cohort. Prev Med 2018;106:45–52. doi: 10.1016/j.ypmed.2017.09.021

10. Chebet JJ, Thomson CA, Kohler LN, et al. Association of Diet Quality and Physical Activity on Obesity-Related Cancer Risk and Mortality in Black Women: Results from the Women’s Health Initiative. Cancer Epidemiol Biomarkers Prev 2020;29(3):591–98. doi: 10.1158/1055-9965.Epi-19-1063 [published Online First: 2020/01/10]

11. Ding D, Van Buskirk J, Nguyen B, et al. Physical activity, diet quality and all-cause cardiovascular disease and cancer mortality: a prospective study of 346 627 UK Biobank participants. Br J Sports Med 2022;56(20):1148. doi: 10.1136/bjsports-2021-105195

12. Williamson EJ, Polak J, Simpson JA, et al. Sustained adherence to a Mediterranean diet and physical activity on all-cause mortality in the Melbourne Collaborative Cohort Study: application of the g-formula. BMC Public Health 2019;19(1):1733. doi: 10.1186/s12889-019-7919-2 [published Online First: 2019/12/28]

13. Alvarez-Alvarez I, de Rojas JP, Fernandez-Montero A, et al. Strong inverse associations of Mediterranean diet, physical activity and their combination with cardiovascular disease: The Seguimiento Universidad de Navarra (SUN) cohort. Eur J Prev Cardiol 2020;25(11):1186–97. doi: 10.1177/2047487318783263

14. Kazemi A, Sasani N, Mokhtari Z, et al. Comparing the risk of cardiovascular diseases and all-cause mortality in four lifestyles with a combination of high/low physical activity and healthy/unhealthy diet: a prospective cohort study. Int J Behav Nutr Phys Act 2022;19(1):138. doi: 10.1186/s12966-022-01374-1

15. Banks E, Redman S, Jorm L, et al. Cohort profile: the 45 and up study. Int J Epidemiol 2008;37(5):941–7. doi: 10.1093/ije/dym184 [published Online First: 2007/09/21]

16. Australian Institute of Health and Welfare (AIHW). The Active Australia Survey: A Guide and Manual for Implementation, Analysis and Reporting. Canberra: AIHW, 2003.

17. Timperio A, Salmon J, Rosenberg M, et al. Do logbooks influence recall of physical activity in validation studies? Med Sci Sports Exerc 2004;36(7):1181–6. doi: 10.1249/01.mss.0000132268.74992.d8 [published Online First: 2004/07/06]

18. Brown WJ, Burton NW, Marshall AL, et al. Reliability and validity of a modified self-administered version of the Active Australia physical activity survey in a sample of mid-age women. Aust N Z J Public Health 2008;32(6):535–41. doi: 10.1111/j.1753-6405.2008.00305.x [published Online First: 2008/12/17]

19. Brown W, Bauman A, Chey T, et al. Comparison of surveys used to measure physical activity. Aust N Z J Public Health 2004;28(2):128–34. doi: 10.1111/j.1467-842x.2004.tb00925.x [published Online First: 2004/07/06]

20. Gebel K, Ding D, Chey T, et al. Effect of Moderate to Vigorous Physical Activity on All-Cause Mortality in Middle-aged and Older Australians. JAMA Intern Med 2015;175(6):970–7. doi: 10.1001/jamainternmed.2015.0541 [published Online First: 2015/04/07]

21. Wang Y, Nie J, Ferrari G, et al. Association of Physical Activity Intensity With Mortality: A National Cohort Study of 403 681 US Adults. JAMA Intern Med 2021;181(2):203–11. doi: 10.1001/jamainternmed.2020.6331

22. Ding D, Rogers K, van der Ploeg H, et al. Traditional and emerging lifestyle risk behaviors and all-cause mortality in middle-aged and older adults: Evidence from a large population-based australian cohort. PLoS Med 2015;12(12):e1001917. doi: 10.1371/journal.pmed.1001917

23. Australian Government Department of Health and Ageing. Eat for health: Australian Dietary Guidelines. Canberra City, ACT: National Health and Medical Research Council 2013.

24. Heart Foundation (2019 update). Eating for Heart Health: Position Statement. Melbourne: National Heart Foundation Australia.

25. Bosco E, Hsueh L, McConeghy KW, et al. Major adverse cardiovascular event definitions used in observational analysis of administrative databases: a systematic review. BMC Med Res Methodol 2021;21(1):241. doi: 10.1186/s12874-021-01440-5

26. Kraus WE, Powell KE, Haskell WL, et al. Physical Activity, All-Cause and Cardiovascular Mortality, and Cardiovascular Disease. Med Sci Sports Exerc 2019;51(6):1270–81. doi: 10.1249/mss.0000000000001939

27. Textor J, van der Zander B, Gilthorpe MS, et al. Robust causal inference using directed acyclic graphs: the R package ‘dagitty’. Int J Epidemiol 2016;45(6):1887–94. doi: 10.1093/ije/dyw341 [published Online First: 2017/01/17]

28. Australian Bureau of Statistics. Census of Population and Housing: Socio-Economic Indexes for Areas (SEIFA), Australia. Canberra, Australia: Australian Bureau of Statistics, 2006.

29. Australian Centre for Housing Research; The University of Adelaide. Accessibility/Remoteness Index of Australia (ARIA+), 2021.

30. Thiébaut AC, Bénichou J. Choice of time-scale in Cox’s model analysis of epidemiologic cohort data: a simulation study. Stat Med 2004;23(24):3803–20. doi: 10.1002/sim.2098

31. Robins JM, Finkelstein DM. Correcting for noncompliance and dependent censoring in an AIDS Clinical Trial with inverse probability of censoring weighted (IPCW) log-rank tests. Biometrics 2000;56(3):779–88. doi: 10.1111/j.0006-341x.2000.00779.x

32. White IR, Royston P, Wood AM. Multiple imputation using chained equations: Issues and guidance for practice. Stat Med 2011;30(4):377–99. doi: 10.1002/sim.4067

33. Association. AH. The American Heart Association Diet and Lifestyle Recommendations 2021 [cited 2023 Aug 9]. Available from: https://www.heart.org/en/healthy-living/healthy-eating/eat-smart/nutrition-basics/aha-diet-and-lifestyle-recommendations.

34. Bull FC, Saad Al-Ansari S, Biddle S, et al. World Health Organization 2020 Guidelines on Physical Activity and Sedentary Behaviour. Br J Sports Med 2020;54:24. doi: 10.1136/bjsports-2020-102955

35. 2018 Physical Activity Guidelines Advisory Committee. 2018 Physical Activity Guidelines Advisory Committee Scientific Report. Washington, DC: U.S. Department of Health and Human Services, 2018.

36. Leandro G, Matthew P, Ali A, et al. Non-occupational physical activity and risk of cardiovascular disease, cancer and mortality outcomes: a dose–response meta-analysis of large prospective studies. Br J Sports Med 2023;57(15):979. doi: 10.1136/bjsports-2022-105669

37. Joshy G, Arora M, Korda RJ, et al. Is poor oral health a risk marker for incident cardiovascular disease hospitalisation and all-cause mortality? Findings from 172 630 participants from the prospective 45 and Up Study. BMJ open 2016;6(8):e012386. doi: 10.1136/bmjopen-2016-012386

38. Brlek A, Gregorič M. Diet quality indices and their associations with all-cause mortality, CVD and type 2 diabetes mellitus: an umbrella review. Br J Nutr 2023;130(4):709–18. doi: 10.1017/s0007114522003701 [published Online First: 20221125]

39. English LK, Ard JD, Bailey RL, et al. Evaluation of Dietary Patterns and All-Cause Mortality: A Systematic Review. JAMA Network Open 2021;4(8):e2122277–e77. doi: 10.1001/jamanetworkopen.2021.22277

40. Paeratakul S, Popkin BM, Kohlmeier L, et al. Measurement error in dietary data: implications for the epidemiologic study of the diet-disease relationship. Eur J Clin Nutr 1998;52(10):722–7. doi: 10.1038/sj.ejcn.1600633 [published Online First: 1998/11/07]

41. Morze J, Danielewicz A, Hoffmann G, et al. Diet Quality as Assessed by the Healthy Eating Index, Alternate Healthy Eating Index, Dietary Approaches to Stop Hypertension Score, and Health Outcomes: A Second Update of a Systematic Review and Meta-Analysis of Cohort Studies. Journal of the Academy of Nutrition and Dietetics 2020;120(12):1998–2031.e15. doi: 10.1016/j.jand.2020.08.076

42. Yuan C, Spiegelman D, Rimm EB, et al. Relative Validity of Nutrient Intakes Assessed by Questionnaire, 24-Hour Recalls, and Diet Records as Compared With Urinary Recovery and Plasma Concentration Biomarkers: Findings for Women. Am J Epidemiol 2018;187(5):1051–63. doi: 10.1093/aje/kwx328

43. Ding D, Do A, Schmidt HM, et al. A widening gap? Changes in multiple lifestyle risk Behaviours by socioeconomic status in New South Wales, Australia, 2002-2012. PLoS One 2015;10(8):e0135338. doi: 10.1371/journal.pone.0135338 [published Online First: 2015/08/21]

44. Mealing N, Banks E, Jorm L, et al. Investigation of relative risk estimates from studies of the same population with contrasting response rates and designs. BMC Med Res Methodol 2010;10(1):26.

45. Bauman A, Lee K, Ding D, et al. Physical activity research in the first 15 years of the “45 and Up” cohort study: a narrative review and citation analysis. Public Health Research & Practice

